# Modification of the Rheumatoid Arthritis (RA) Patient Reported Experience Measure (PREM) for Patients with Interstitial Lung Disease (ILD)

**DOI:** 10.1101/2025.04.09.25325349

**Authors:** J Mandizha, C Crook, J Lanario, R Davies, A Duckworth, AP Howard, S Lines, M Gibbons, C Scotton, AM Russell

## Abstract

**Objective:** Patient reported experience measures (PREMs) are a key component of healthcare accountability frameworks, health policy, integrated care board (ICB) commissioning and integrated care partnerships (ICPs) generating data which are crucial markers of patient care quality. The Rheumatoid Arthritis Patient Reported Experience Measure (RA-PREM) incorporates the eight core elements of NHS Patient Experience Framework and is validated in a range of rheumatic conditions. Our objective is to determine the acceptability and feasibility of the RA-PREM for an ILD population.

**Design:** A mixed-methods patient-centred approach incorporating an interdisciplinary research steering group with patient-partners. Patient surveys evaluated the language and meaning of the RA-PREM eight domains, twenty-four statements and response categories. A patient focus group examined contentious statements. A consensus group of expert patient-partners agreed statements for the modified RA-PREM. Focus group participants reviewed the modified instrument (ILD-PREM) for acceptability and face/content validity.

**Setting:** A single NHSE-commissioned, regional ILD service/UK.

**Results:** Thirteen patients (10 male) diagnosed with ILD participated in focus group discussions. Critical discussion of the RA-PREM resulted in nuanced modifications of four statements of three domains. Five patients (3 male) and three healthcare researchers attained consensus on the face/content validity of statements. Seventy-three patients completed the ILD-PREM following outpatient contact.

**Conclusion:** The ILD-PREM retains twenty-four statements representing the eight domains of the RA-PREM. It meets face/content validity criteria and is acceptable to an ILD population. Longitudinal validation of the ILD-PREM across ILD services including further testing in global minority groups will establish criterion and construct validity and objective measures of reliability.

**Key Messages:**

- *What is already known on this topic*
  - Patient experience metrics are a key component of healthcare quality frameworks.
  - Better patient experience is associated with better quality of care, higher treatment adherence and improved patient outcomes.
- *What this study adds*
  - The RA-PREM has been adapted for use within ILD, using patient-centred methods.
  - The ILD-PREM retains twenty-four statements capturing level of agreement which map to the eight indicators of the NHS Patient Experience Framework.
  - The ILD-PREM has face/content validity and clinical utility for an ILD population.
- *How this study might affect research, practice, or policy*
  - This is the first disease-specific PREM for ILD enabling data collection for service evaluation and improvement.
  - It is judicious to modify validated measures to obviate the need for resource intensive development of a new instruments where possible.

## Introduction

Interstitial Lung Diseases (ILD’s) are characterised by irreversible inflammation and scarring of the lung tissue that is often progressive. Associated with high morbidity and mortality rates, symptoms of ILD include debilitating breathlessness, cough and fatigue^1,2^. ILD is the primary pulmonary manifestation in Rheumatoid arthritis (RA) and connective tissue diseases (CTDs), such as scleroderma (systemic sclerosis), inflammatory myositis (polymyositis and dermatomyositis), Sjögren syndrome, and undifferentiated CTD^3^. Symptomology is often shared and optimal healthcare requires palliation, slowing disease progression and improving or maintaining quality-of-life (QoL). The efficacy of this approach can be measured by clinical biomarkers and the metrics of serial data of patient-reported measures (PRMs). PRMs guide decisions in palliative care such as outcomes, lived experience, motivation or treatment preference.

The National Institute for Health and Care Excellence (NICE) Quality Standard, endorsed by NHS England, recommends follow-up appointments for patients with Idiopathic Pulmonary Fibrosis (IPF)^4^, every 3-12 months, adjusted according to disease stability^4,5^. This standard for IPF, the most common form of ILD, may be extrapolated to all ILDs. The clinical review minimally includes physical assessment and pulmonary function tests with the opportunity for oxygen assessment, appropriate onward referral for pulmonary rehabilitation, lung transplantation or palliative care^4^. Patient Reported Outcome Measures (PROMs) integrated in the clinical review gather self-reports of symptoms, impact of symptoms and psychosocial status providing essential metrics and assisting clinical decision-making. Three key objectives of health and care activities are their direct benefit to patients, such as improved health outcomes; how patients and their family and friends experience healthcare episodes, and the effective use of financial resources. While individuals have professional responsibility to maintain and improve quality of care, healthcare quality frameworks offer population oversights that identify health inequalities with potential to improve health outcomes^6^.

The metrics of patient experience underpinned by a philosophy of patient-centred care influences the decision-making of healthcare providers, integrated care boards (ICB) and key stakeholders’ further impacting service provision^7^. We cannot improve aspects of health services if we do not collect and evaluate these metrics. The NHS Patient Experience Framework has identified eight indicators to guide measurement of what is reported to be the most important elements of patient’s healthcare experiences^8–9^. See Table 1.

**Table 1:** Eight indicators of the NHS Patient Experience Framework.

|  |  |
| --- | --- |
| Respect for patient-centred values & preferences | Welcome the involvement of family and friends |
| Coordination and integration of care | Information, communication & education |
| Emotional support | Transition and continuity |
| Physical comfort | Access to care |

### Properties of Patient Reported Experience Measures

Patient-Reported Experience Measures (PREMs) use Likert scale scoring to quantify a range of care quality, including relational and functional elements of healthcare experience. The Relational PREM statements focus on the patient-clinician relationship while functional PREM statements focus on the objective experiences of care facilities, for example, access, cleanliness, or comfort. PREMs, like the other PRMs may be generic or disease specific^10^ and may have utility across a range of health and social care settings.

Three systematic reviews (SRs) examined the psychometric testing of PREMs. The first in a mixed inpatient population^10^, the second in emergency care service provision^11^ and the third across a range of health care contexts^12^. Beatie et al^10^ identified 26 papers for inclusion including 11 international instruments. Authors report evidence of extensive theoretical/development work and whilst the quality of methods and results were variable mostly the standard was high. The PREMs included were acceptable to context but the evidence on the ease of use was poorly reported for over half of the measures. Male et al^11^ identified 8 papers measuring patient experience of emergency hospital care including 4 international instruments. Similarly, content validity & theoretical development was well reported. Their evaluation concluded that the validity and reliability of the PREMs should be reassessed throughout development. Although none of the studies reported on responsiveness the PREMs for use within the ED identified in this SR were found to be adequately developed. Bull et al^12^ identified 88 PREMs in 109 papers meeting inclusion criteria across four main health care contexts of inpatient care services (36.4%), primary care (23.9%) outpatient care services (12.5%). Fifty-nine of the PREMs were generic measures. The PREMS were evaluated using the COSMIN framework reporting on validity & reliability, specifically Internal consistency structural & content validity most oft reported. Overall greater rigour was needed in the psychometric evaluation of each PREM^13^.

Given the number of PREM instruments in circulation we were interested in those used or developed for use in conditions clinically associated with ILD. The Hospital Consumer Assessment of Healthcare Providers and Systems (HCAHPS)^14^, a 29-item generic PREM specifically for inpatient care, was reported to have utility in patients with chronic obstructive pulmonary disease (COPD) admitted with acute exacerbations^10^. The HCAHPS was the preferred PREM in the CICERO–ELF patient survey of 200 participants^15^. The HCAHPS is included in the Hospital Value-Based Purchasing program to calculate value-based incentive payments in the US and is focused on the perspective and experiences of hospital admissions.

The PREM-C9, a COPD-specific PREM captures patients experiences and interactions with healthcare systems and clinicians in three areas of healthcare: ‘usual care’ (items 1-3); ‘everyday day life’ (items 4-7); and ‘self-management and exacerbations’ (items 8-9). Assessed for validity and reliability and expanding on the psychometric testing of the original UK PREM-C9 English version the Catalan and Spanish versions of the PREM-C9 showed good reliability and known groups’ validity suggesting that the PREM-C9 is appropriate for use as a measure of overall COPD experience using the overall score^16^. Although translated in other languages use of the PREM-C9 is only reported in COPD to measure the domains of everyday life; usual care and exacerbations across the spectrum of severity of disease.

We considered the merits of the existing PREMS both generic and disease specific and whether there was indeed a need to develop a new instrument specifically for IPF/ILD. The lack of appropriate PRMs including PREMs in IPF has contributed to gaps in the field^17,18^ and currently, there are no PREMs available for clinical use^19^. The Patient Experiences and Satisfaction with Medication Questionnaire (PESaM) may have some utility on medication experiences (specifically Pirfenidone) in IPF.^20^

The Rheumatoid Arthritis Patient Reported Experience Measure (RA-PREM) developed by the CQRA (Commissioning for Quality in Rheumatoid Arthritis) used patient-centred methods^21^. Focus groups identified key elements of the patient experience which researchers mapped to the NHS framework^8^. The Cronbach’s alpha ranged from 0.76 to 0.91 across its eight domains demonstrating good internal consistency of the RA-PREM in 524 patients across 10 UK sites. The RA-PREM validated in the original study^21^ has subsequently been validated in other global rheumatology populations demonstrating feasibility, acceptability, relevance, validity and reliability^22–23^.

The two candidate instruments for the adaptation for use in an ILD population, the PREM-C9 and RA-PREM were evaluated by our interdisciplinary research group. A preference for the RA-PREM was determined as it offered standard closed-ended questions thought offer time saving and useful data while a comments section offered narrative information with ‘supplemental value’ on detail and context. The RA-PREM was determined to be culturally and clinically relevant and we were granted permissions to adapt the RA-PREM for an ILD population.

## Aims

- *To establish acceptability and feasibility of modification of the RA-PREM for an ILD population*.
- *To modify the content and language of the RA-PREM as indicated using patient centred and consensus methods*.
- *To produce an acceptable PREM for validation in an ILD population (ILD-PREM)*.

## Methods

### See the infographic of the methodological approach in Figure 1

#### Feasibility

Patient centred researchers (JM, AD, AMR) convened an interdisciplinary ILD steering group of patient-partners and clinicians (n=10) to discuss the feasibility and utility of the RA-PREM for an ILD population and determine the need for modification.

**Figure 1:**
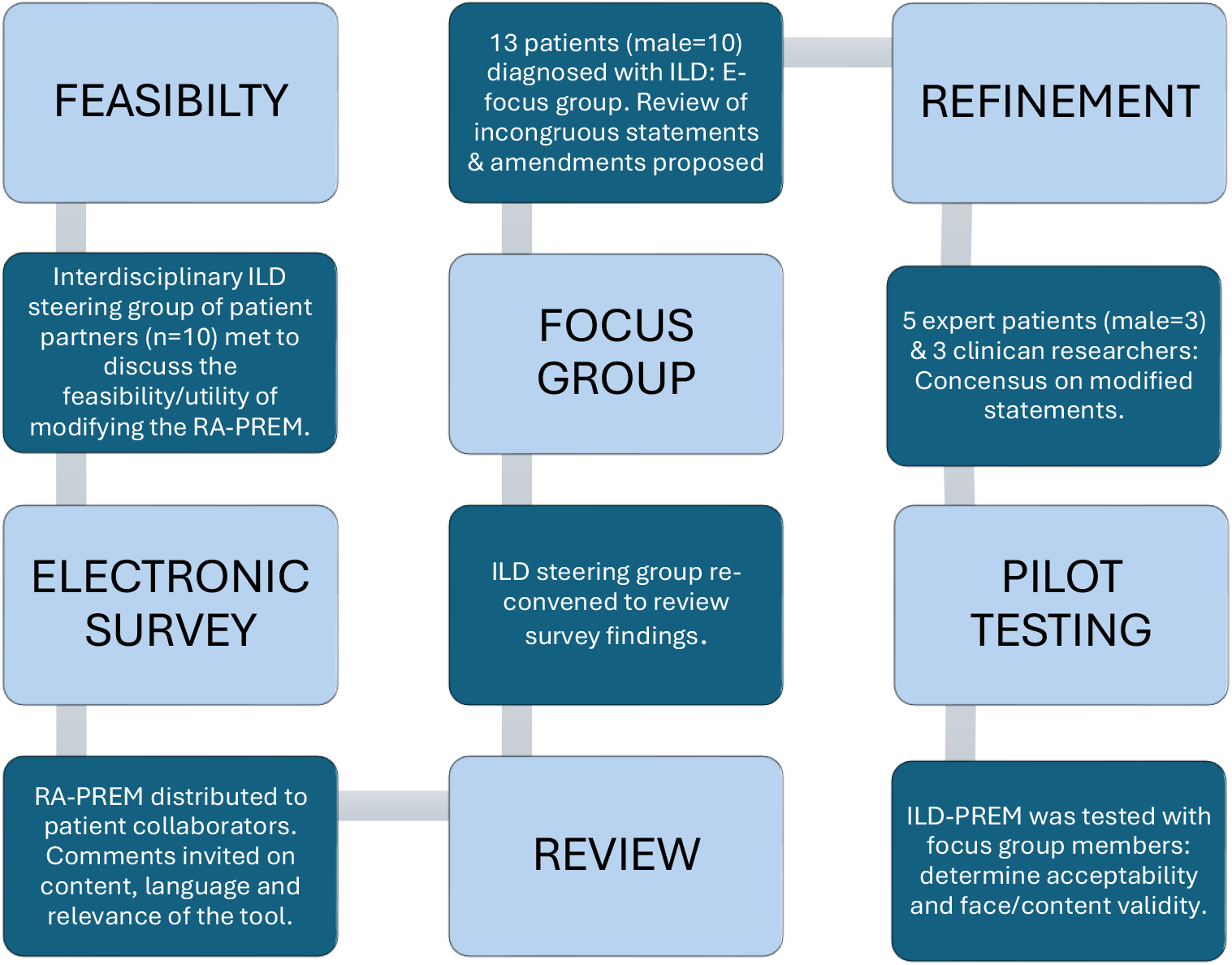
Methodological Approach.

#### Electronic Survey

The research team adapted an electronic version of the RA-PREM, changing explicit references to RA disease to lung disease and distributed it to members of the ‘Exeter Patients in Collaboration for Pulmonary Fibrosis Research’ (EPIC-PF) group of patients diagnosed with ILD across a range of severity and trained in critical appraisal of research (n-14). EPIC members agreed to comment on the content, language, and relevance of each item. Subsequently the EPIC group convened a meeting to review comments.

#### Focus Group

Thirteen patients (male=10) diagnosed with ILD participated in a 2-hour online focus group to discuss all statements of the RA-PREM. Each statement was read in turn, and comments were invited as to relevance, perceived meaning and need for modification. Discussion focussed on incongruous statements and the proposal of amendments. Two female 4researchers (JM; AMR) experienced in qualitative research methods and analysis, facilitated the audio recorded group and kept field notes. We did not generate transcripts, rather listened and relistened to the recording to augment field notes and identify items that required modification.5

#### Refinement

Four expert patients (male=3) who had an established working relationship with the research team, participated in cognitive interviews to achieve consensus of the meaning of modified items via an electronic platform. The penultimate version of the modified PREM (ILD-PREM) was circulated to the EPIC group and research team for further comment or final approval.

#### Pilot Testing

Having agreed the final ILD-PREM, focus group members reviewed its’ acceptability and face/content validity. A pilot study evaluated the acceptability of the ILD-PREM at a single-centre, specialist ILD service. The ILD-PREM was sent to all newly referred patients attending outpatient clinic over a six-month period (August 2023 to February 2024) using a consecutive sampling approach. At the consultation, the researchers gave consenting patients a paper copy of the ILD-PREM with. instruction on how to complete it and how to return the form anonymously in a pre-paid envelope within two weeks of their appointment.

#### Patient and Public Involvement

Patient involvement was integral to this work and, patient-partners were involved from initial conversations about feasibility, through to focus group participation, refinement, and pilot testing. The EPIC group are an established group of patient partners in research.

#### Statistical analysis

Cronbach’s alpha calculated the internal consistency of the ILD-PREM for twenty qualifying questions. Questions 7b and 7d require binary response options and participants only complete questions 7c and 7e if they answer “yes” to 7b and 7d. We excluded these four questions from the Cronbach’s alpha calculation as they did not meet all the assumptions of this statistical test.

## Results

The interdisciplinary ILD steering group (n=10) unanimously agreed the appropriateness of the RA-PREM, its robustness and usefulness in modification.

The EPIC group members (n=10) reviewed the electronic RA-PREM commenting on each item. In turn the steering group members examined each item and response. Where most comments were positive or neutral *(‘Clear and easy to answer,’ ‘pertinent and phrased well,’ ‘highly relevant-well put’ ‘No comment’*), the original statements were retained.

Four statements consistently generated critical comments. Criticism ranged from being purely practical, for example, one statement referenced ‘attending clinic’ and respondents identified that clinics are now often virtual. They considered other statements to be irrelevant or poorly worded in the context of ILD. For example, the statement ‘I feel that my lung condition is being controlled enough to let me get on with my daily life and usual activities’ generated the following responses: *‘This is not relevant for a progressive condition,’ ‘This is a bit vague! Not my usual activities before I had IPF’ ‘Not controlled enough could either be the nature of the condition or the support being given*.*’*

The steering group convened a focus group to examine these four statements. Focus group members proposed modification and alternative content or language for the four statements. One participant commented on the possibility of *‘control’* of symptoms of diseases such as Idiopathic Pulmonary Fibrosis (IPF) *‘no amount of treatment will help what I have*.*’*

There was recognition that ‘daily life’ or ‘usual activities’ vary, and the presence of a progressive and irreversible lung disease may permanently alter functioning. *‘Adapting’* to what our patient group described as this *‘new normal’* was a key component, alongside recognising that change was inevitable within this disease context.

Post Focus group, researchers (JM; AMR) compared field notes and redrafted the statements accordingly. We circulated the revised statements to the expert patient group, for refinement and clarification of meaning and consensus discussion. See table 2.

**Table 2:** Four Amended Statements of the RA-PREM.

| Domain 1: Your needs and preferences |  |
| --- | --- |
| **Original Statement** | *Whenever I attended clinic, I felt that I was treated respectfully as an individual* |
| **Modified Statement** | *During my appointments, I felt that I was treated respectfully as an individual* |
| Domain 4: Daily living and physical comfort |  |
| **Original Statement** | *If I have had an exacerbation (when my symptoms got much worse), I have been able to get help quickly* |
| **Modified Statement** | *If my symptoms get much worse (e.g. I experience an exacerbation or flare), I have been able to get help quickly* |
| Domain 4: Daily living and physical comfort |  |
| **Original Statement** | *I feel that my lung condition is being controlled enough to let me get on with my daily life and usual activities* |
| **Modified Statement** | *I feel that I have the right support to adapt and manage my daily activities around my changing lung condition* |
| Domain 5: Emotional support |  |
| **Original Statement** | *I feel able to discuss personal or intimate issues about relationships with my health team if I want to* |
| **Modified Statement** | *I feel able to talk about personal or intimate issues with my health team, if I want to* |

The final version of the questionnaire was circulated to the expert patient group EPIC-PF, who agreed that the questionnaire was now appropriate for use in an ILD population, being *‘clear, focused, well laid out and easy to understand*.*’*

We evaluated the ILD-PREM at a regional specialist ILD centre covering a diverse geographical catchment area in the Southwest of England. One hundred and forty-eight patients attended twenty-eight clinics during the 6-month test period (137 face-to-face, eleven via telephone). Patients had waited for their initial assessment an average of 5 months, following referral (range: 1-10 months).

One hundred and forty-eight patients agreed to complete the questionnaires with seventy-three returned (response rate 49.3%). Thirty-six respondents were male, twenty-one identified as female with sixteen respondents not declared. While the most common ILD diagnosis, IPF was the most disproportionately represented diagnosis (n=51), and it was therefore unsurprising that with more than half of respondents were in the age range 70-79 years. See Table 3.

**Table 3:** Characteristics of respondents.

| Characteristics | Cohort | Respondents |
| --- | --- | --- |
| **Total numbers (n)** | 128 | 73 |
| **Gender, n (%)** |  |  |
| Male |  | 36 (49.3) |
| Female |  | 21 (28.8) |
| Non-Binary |  | 0 (0.0) |
| Prefer not to say |  | 1 (1.4) |
| Not Answered |  | 15 (20.5) |
| **Age (years), n (%)** |  |  |
| 39 or less |  | 0 (0.0) |
| 40 to 49 |  | 1 (1.4) |
| 50 to 59 |  | 2 (2.7) |
| 60 to 69 |  | 18 (24.7) |
| 70 to 79 |  | 39 (53.4) |
| 80 to 89 |  | 9 (12.3) |
| 90 and over |  | 0 (0.0) |
| Not Answered |  | 4 (5.5) |
| **Ethnicity, n (%)** |  |  |
| White (British/Irish/Other white background) |  | 70 (95.9) |
| Mixed (White & Black Caribbean or African/White & Asian/Other mixed background) |  | 0 |
| Asian / British Asian (Indian/Pakistani/Bangladeshi/Other Asian) |  | 0 |
| Black or Black British (Caribbean/African/Other Black) |  | 0 |
| Chinese |  | 0 |
| Other Ethnic Group |  | 0 |
| Prefer not to say |  | 0 |
| Not Answered |  | 3 (4.1) |
| **Diagnosis, n** |  |  |
| Idiopathic Pulmonary Fibrosis | 77 | 51 |
| Hypersensitivity Pneumonitis | 27 | 9 |
| Connective Tissue Disease associated ILD | 3 | 3 |
| Unclassifiable ILD | 4 | 10 |
| Don't know/Diagnosis uncertain | N/A | 1 |
| Other (please specify) | 37 | 1 |
| Not Answered | N/A | 6 |
| **Years since diagnosis, n (%)** |  |  |
| Less than 6 months |  | 7 (9.6) |
| Between 6 months and 1 year |  | 18 (24.6) |
| 1 to 2 years |  | 14 (19.2) |
| 2 to 3 years | N/A | 7 (9.6) |
| More than 3 years |  | 21 (28.8) |
| Not Answered |  | 6 (8.2) |
| **Forced Vital Capacity (FVC) % predicted, n** |  |  |
| 30 – 50% | 16 |  |
| 51 – 70% | 54 |  |
| 71 – 90% | 48 | N/A |
| 91 – 130% | 29 |  |
| Not available | 1 |  |

Response option frequencies are available in the supplementary file Table 4: Distribution of ILD PREM responses. We note the data is positively skewed, but do not make inferences, as there is no comparative measure.

Of the seventy-three questionnaires returned, fifty-nine statements contained ‘no answer’ response. These missing data equate to 3.4% of the total number of questionnaires returned. Questions most frequently not answered were:

- *7d) I have needed extra treatment or a change of treatment (between routine clinic appointments)* (n=9 respondents). Inference: This may relate to the fact that this was patients’ first appointment.
- *4b) If my symptoms get much worse (e*.*g. I experience an exacerbation or flare), I have been able to get help quickly* (n=9 respondents). Inference: It is possible that the respondents may not have experienced an exacerbation.
- *7b) I have had appointments cancelled unexpectedly* (n=7 respondents)
- *5a) I feel able to approach a member of my health team to discuss any worries about my condition and my treatment or their effect on my life* (n=5 respondents)

These questions all appear on the second page of the questionnaire and we hypothesis a proportion of the respondents may have overlooked the page. Given the commitment to anonymity, we are unable to definitively establish the reasons for the unanswered questions. Evidence for structural validity of a scale is a prerequisite for interpretation of internal consistency analysis, a measure of internal structure of an instrument. Cronbach’s α ≥0.70 is considered indicative of sound reliability for subscale items, as well as overall scores. The Cronbach’s α was 0.78 for total scores of completed ILD-PREM questionnaires (n= 51), excluding the 4 questions listed above from this analysis and indicating a good internal consistency for the overall scale.

## Discussion

Using patient-centred approaches we have modified the RA-PREM^21^ to ensure the language and content was relevant to an ILD population. The twenty-four statements retained have linkage to the eight indicators within the NHS Patient Experience Framework^8^.

Two statements, relating to Domains 1 and 5, required only minor changes following patient feedback. Two statements, both linked to Domain 4 (Daily Living and Physical Comfort) needed substantial rewording, not surprisingly in the context of ILD. The RA-PREM statement refers to a ‘flare’, which is a concept well understood in Rheumatology. Albeit a contentious issue within ILD, being poorly defined and lacking clinician consensus and standardised patient understanding we changed ‘flare’ to the more widely used term ‘exacerbation.’ Patients with lived experience perceived this broader definition of decline to encapsulate ‘symptoms getting much worse.’

Likewise, within the next statement, ‘control’ is an appropriate treatment goal for rheumatological conditions, but not a realistic aim for the insidious, complex, and frequently progressive symptoms associated with ILD, particularly breathlessness. The result was a much more nuanced statement, referring to support in adapting and managing daily activities. The modified statement recognises that change is inevitable, thereby removing the word ‘usual’, which is unhelpful. Full ‘control’ is unrealistic, and the passive statement (‘being controlled’) becomes an active one.

Our work demonstrates that modifying validated measures in this way, where there is reasonable homogeneity in disease course and symptom burden is a feasible and efficient way of developing measures for a different patient group. This approach is efficient and maximises the original research investment of time and resource. The work demonstrates the importance of recognising the complexities relating to chronic disease and the value of disease-specific measures^16,19,21^, as well as the importance of patient involvement in the development of measures^24^.

Recent systematic reviews have highlighted complex unmet needs in this patient population including a lack of psychological support throughout the course of living with pulmonary fibrosis^25,26^. Misconceptions and barriers to the provision of palliative care and a gap between informational needs and provision prevail^26^. The introduction of oxygen is a significant event both psychologically and domestically^25^. Collecting PREM data at each encounter will offer insight at a population and service level informing targeted interventions to improve and reduce the burden of unmet needs.

Patients offer a unique perspective on the relevance and language of questionnaire content, helping to ensure tools accurately measure what we think we are measuring increasing the quality of our data (face and content validity). While this pilot testing of the ILD-PREM indicates acceptability of the measure within a specific population, further work will assess the acceptability of the ILD-PREM in more diverse groups. This includes patients from global majority ethnicities underrepresented in our data but also includes patients with a wider range of ILD diagnoses. IPF is the most common form of ILD, accounting for 20-50% of cases^2^. Our demographic data show a disproportionate representation, at 70%. Further study will explore validity and additional reliability testing to establish the ILD-PREM appropriately captures patient-reported experiences of health care across the range of ILDs.

All patient reported measures including PREMs, have limited usefulness in the absence of data. Missing data often results from non-responders, including those with low literacy levels, limited English, or poor health. We distributed the ILD-PREM in paper format, but future work will likely require a hybrid approach with the option for an e-PREM. Exploring non-response patterns was beyond the scope of this preliminary work, however, this will be important in future work to address escalating health inequalities

## Conclusion

PREMs are useful tools to monitor and, subsequently, improve patient experience of care. Using patient-centred methods we have modified the RA-PREM for use in an ILD population. The ILD-PREM retains twenty-four statements, which are representative of the eight indicators the PREM seeks to measure. Our expert patient group has confirmed face and content validity of the ILD-PREM. Participants in the pilot study confirmed acceptability of the ILD-PREM. Longitudinal studies will provide a larger data set to enable further psychometric testing using exploratory factor analysis and calculations of median time of completion to ensure that time constraints is not a barrier to completing the ILD-PREM. Objectively measuring patient experience within the wider ILD community will add to the quality of clinical care.

## Data Availability

All relevant anonymised data produced in the present work are contained in the manuscript. Additional data produced in the present study, i.e. transcribed audio recordings redacted for anonymity are available upon reasonable request to the authors

## Acknowledgement

We are grateful to Mr Graham Brown and Carol Bond for their altruistic work in supporting this project. Both were committed to enhancing the patient experience of those living with Interstitial Lung Diseases. Sadly, Graham and Carol passed away prior to publication, a stark reminder of the devasting mortality associated with ILDs. They remain in our thoughts and our work.

## Ethics

Institutional ethical approvals for the study were granted by the Royal Devon University Hospitals NHS Trust (RDUHT) Independent Respiratory Speciality Governance committee and accordingly registered as a Quality Improvement project reference number 24-1413. The body is accountable to the director of governance Melanie Holley and the RDUHT Board of directors. The independent panel members who reviewed this protocol: Mr Jacob Bruun (RDUHT Lead for Safety & Quality Improvement), Dr Nicholas Withers (Consultant Respiratory Medicine; lead for adult clinical CF services RDUHT & lead for CF Trust research trials); Dr Thomas Burden (Consultant Respiratory Physician, Clinical lead for Respiratory services RDUHT & fellow NIHR University of Exeter BRC).

## Author Contributions

The authors confirm contribution to the manuscript as follows: AMR is the guarantor; study conception and design: AMR JM; data collection: JM, CC, RD; Analysis and interpretation of the results: AMR JM JL AD RD CC; Draft manuscript preparation: JM AMR JL AD RD HA SL CS. All authors reviewed the results and approved the last version of the manuscript.

## Funding

This work was supported by a southwest CRN internship award

## Patient and public involvement

Patients engaged in the design, conduct, reporting, and dissemination plans of this work.

## Conflict of Interest

JM is supported by NIHR-CRN funding to undertake this work; JL is in receipt of grants from European Respiratory Society, AstraZeneca and GlaxoSmithKline. AMR received speaker fees from Boehringer-Ingelheim and an international fellowship award from the University of

Adelaide, Australia.

## Notes

### Funding Statement

JM was funded by the Peninsular NIHR CRN whilst conducting this work

### Author Declarations

Institutional ethical approvals for the study were granted by the Royal Devon University Hospitals NHS Trust (RDUHT) Independent Respiratory Speciality Governance Committee and accordingly registered as a Quality Improvement project reference number 24-1413. The body is accountable to the director of governance Melanie Holley and the RDUHT Board of directors. The independent panel members who reviewed this protocol: Mr Jacob Bruun (RDUHT Lead for Safety & Quality Improvement), Dr Nicholas Withers (Consultant Respiratory Medicine; lead for adult clinical CF services RDUHT & lead for CF Trust research trials); Dr Thomas Burden (Consultant Respiratory Physician, Clinical lead for Respiratory services RDUHT & fellow NIHR University of Exeter BRC).

